# Protocol for evaluation of sustained virological response as a surrogate outcome for mortality, decompensated cirrhosis, or hepatocellular carcinoma in people with chronic hepatitis C virus infection treated with direct-acting antivirals

**DOI:** 10.1101/2024.10.22.24315958

**Authors:** Kurinchi Gurusamy, Christian Gluud

## Abstract

**Introduction:** Sustained virological response (SVR) is commonly used as an indicator of treatment success in people with chronic hepatitis C virus (HCV) infection. However, there is uncertainty on whether SVR is a validated surrogate marker of successful treatment of chronic HCV infection.

**Aim:** To evaluate whether SVR is a good surrogate for all-cause mortality, decompensated cirrhosis, or any specific aspect of liver decompensation (jaundice, ascites, hepatic encephalopathy, hepatorenal syndrome, or variceal haemorrhage), or hepatocellular carcinoma in people with chronic HCV infection eligible to receive direct-acting antiviral drugs.

**Methods:** *Data source:* Two ongoing systematic reviews on the effectiveness of direct-acting antiviral drugs in chronic HCV infection.

*Analysis:*

1. Estimate the regression coefficients or between-studies correlation between SVR and the event by three different Bayesian approaches with OpenBUGS, as outlined in the guidance by the Evidence Synthesis Unit (Technical support document 20).
2. Estimate the average proportion of the effect mediated through SVR by causal mediation analysis using R.

**Discussion:** We will use the German Institute of Quality and Efficiency in Health Care (IQWiG) criterion for surrogacy for cancer and at least 50% of the treatment effect mediated through SVR but will report the information in a way that allows people to interpret the information using their own criteria.

## Introduction

Sustained virological response (SVR) is commonly used as an indicator of treatment success in people with chronic hepatitis C virus (HCV) infection. In a Cochrane systematic review, of the 138 trials included, only 11 trials reported all-cause mortality while HCV-related morbidity such as decompensated cirrhosis was not reported in any of the trials (1). On the other hand, 32 trials reported on the surrogate outcome sustained virological response (SVR) (1). Some consider that SVR is the fundamental goal in treatment of HCV (2), although convincing valid proof of this surrogate for clinical important outcomes has been hard to identify (1). On the other hand, others dispute whether SVR is a validated surrogate marker of successful treatment of chronic HCV infection (1,3-6).

## Aim

To evaluate whether SVR is a good surrogate for all-cause mortality, decompensated cirrhosis, or any specific aspect of liver decompensation (jaundice, ascites, hepatic encephalopathy, hepatorenal syndrome, or variceal haemorrhage) (7), or hepatocellular carcinoma in people with chronic HCV infection eligible to receive direct-acting antiviral drugs.

## Methods

### Data source

The Cochrane systematic review on the effectiveness of direct-acting antiviral drugs in chronic HCV infection (1) is currently being updated (personal communication). In addition, a new review on the effectiveness of direct-acting antivirals from observational studies will be conducted (8). As part of these reviews, the information shown in **Error! Reference source not found**. or adjusted treatment effects of the intervention versus comparator on SVR and event will be collected. In the context of this figure and the protocol, an event is one of the following.

1. All-cause mortality
2. Decompensated cirrhosis, i.e. jaundice, ascites, hepatic encephalopathy, hepatorenal syndrome, or variceal haemorrhage.
3. Any specific aspect of liver decompensation (jaundice, ascites, hepatic encephalopathy, hepatorenal syndrome, or variceal haemorrhage)
4. Hepatocellular carcinoma.

### Analysis

For each event, we will evaluate whether SVR is a good surrogate outcome for the clinical event. We will reconstruct the individual participant data from the summary data since this is a binary outcome. We will then perform the following to achieve the research objectives.

1. Estimate the regression coefficients or between-studies correlation between SVR and the event using the guidance by the Evidence Synthesis Unit (Technical support document 20) (9) by three different Bayesian approaches using OpenBugs (10), namely, standard surrogacy model by Daniels and Hughes (11), bivariate random-effects meta-analysis model (9), and bivariate random-effects meta-analysis in product normal formulation model (12). We will use three chains and use non-informative priors. We will start with a burn-in of 10,000 iterations and run further 30,000 iterations to calculate the parameters along with their credible intervals (CrI). We will check for convergence visually and will increase the iterations, use ‘overrelax’ or ‘thin’, as required to achieve convergence. For the treatments effects of SVR and event (clinical outcome) which are required to use these approaches, we will obtain adjusted odds ratio when available. If not available, we will calculate the odds ratios and their 95% confidence intervals (CI) of SVR and event using R (package: meta) (13) from the total number of participants and the number of participants with SVR or events, respectively. If there are zero events, we will use a correction factor of 0.01 to obtain these measures. For estimating the correlation between SVR and the event, we will use the model (among the three models mentioned above) with best (lowest) deviation information criterion to base our conclusions. For the between-study correlations estimated from the bivariate random-effects meta-analysis model (9), and bivariate random-effects meta-analysis in product normal formulation model (12), we will use the German Institute of Quality and Efficiency in Health Care (IQWiG) criterion for surrogacy, which requires a correlation with the lower limit of the 95% confidence interval above 0.85 (14). In fact, we will use 95% CrI as we will use Bayesian methods to indicate that SVR is a good surrogate outcome for the clinical event. If this cannot be achieved, we will calculate the 80% CrI and see if the lower limit of the 80% CrI was above 0.85 (14). If the lower limit of 80% CrI but not the 95% CrI is above 0.85, we will consider that SVR may be a good surrogate outcome for the clinical event, but that there is uncertainty around this conclusion and further studies are necessary. For the standard surrogacy model by Daniels and Hughes, we will consider whether the 95% CrI of the intercept overlaps 0 and the 95% CrI of the association between the treatment differences on SVR and the clinical outcome does not overlap 0 as criteria for SVR being a good surrogate outcome.
2. We will perform a causal mediation analysis and estimate the average causal mediation effects (ACME) and the average direct effects (ADE) using R (package: mediation) (15). We will estimate the average proportion of the effect mediated through SVR and will consider SVR as a surrogate outcome if the lower 95% confidence intervals (CI) of the proportion of the effect mediated through SVR is more than 0.5.

#### Evidence from randomised clinical trials compared to those from observational studies

We will analyse the information from randomised clinical trials separately from those from observational studies. If the conclusions based on observational studies is different from those based on randomised clinical trials, we will highlight this difference and indicate the need for corroborating the information from randomised clinical trials.

### Data sharing and reporting

The summary data will be available from the systematic review update and all authors of the two reviews will be invited to co-author the results of this research. The data and codes used for this research will be shared via www.zenodo.org. Attempts will be made to publish the report in a journal. If this is not possible, we will share the report of our results through www.zenodo.org.

## Discussion

There is considerable controversy about whether SVR is a good surrogate outcome for clinical outcomes in chronic HCV infection (1,3-6). However, there has been no previous meta-study evaluating whether SVR is a good surrogate outcome for clinical outcomes in people eligible to receive direct-acting antivirals in the treatment of chronic HCV infection. We will use two different approaches to perform this evaluation. These approaches have been specifically developed to evaluate whether an outcome is a surrogate outcome across a range of medical conditions or whether the effect on an intervention on an outcome is mediated by a surrogate outcome, which is the rationale behind the use of surrogate outcomes.

In the first approach, for bivariate random-effects meta-analysis model (9) and bivariate random-effects meta-analysis, we will use correlation coefficients to evaluate whether SVR is a good surrogate outcome. There is no consensus on the thresholds used for interpreting correlation coefficients (9). We will use the IQWiG guidance on surrogate outcomes in oncology (14) to interpret whether SVR is a good surrogate outcome. If the lower limit of the 95% CrI overlaps 0.85, we will check whether the lower limit of the 80% CrI overlaps 0.85. Therefore, we will highlight this additional uncertainty.

Although the limit of 0.85 for correlation coefficient has been suggested for cancers, we will use these limits for chronic HCV infection. When there is a high probability of a clinical outcome, one might want to take a more lenient approach to accept surrogate outcomes and treatments that reduce the surrogate outcome, when there is a requirement to act early. In the case of cancer, the proportion of people dying within 10 years is 50% across all cancers (16). Therefore, one might want to take a more lenient approach in approving treatments based on surrogate outcomes. With HCV infection, the proportion of people dying in about 10 years is around 13% (17): this is much less than those dying because of cancers. Therefore, one might argue that much stricter criteria should be used for approving treatments for chronic HCV infection based on surrogate outcomes, if the treatments are funded by the public. Others might argue that because of the longer latency period in chronic HCV infections, there are a lot of factors that might change over time and use this argument to justify a more lenient approach to approving treatments on surrogate outcomes. However, when the main purpose is to maximise the health of a population rather than an individual, the longer period of latency in chronic HCV infections goes against using a more lenient approach (for validating and using a surrogate outcome for funding treatment of chronic HCV infection) in an already stretched healthcare system. To avoid using some new arbitrary threshold criteria, we decided to use similar criteria as used for cancer. Furthermore, we will present the correlation coefficients with the 95% CrI (and possibly 80% CrI) along with the data and the codes used for our analyses. If an individual or an organisation is interested in using a different threshold for interpreting whether SVR is good surrogate outcome for clinical outcomes in people eligible for direct-acting antiviral treatment, they can use the data, codes, and results to guide their decision utilising an alternate threshold.

If the standard surrogacy model by Daniels and Hughes has the best fit, we will consider whether the 95% CrI of the intercept overlaps 0 and the 95% CrI of the association between the treatment differences on SVR and the clinical outcome does not overlap 0 as the criteria for SVR being a good surrogate outcome. This is based on the guidance provided by Daniel and Hughes in interpreting the results (11). The rationale for this approach is as follows. If the entire effect of the treatment on the clinical outcome is mediated through the surrogate outcome, a treatment that does not have any effect on the surrogate outcome must have no effect on the clinical outcome resulting in the intercept being 0 (11). Furthermore, if the CrI of the association between the treatment differences on SVR and the clinical outcome overlaps 0, this indicates that there is a possibility that there is no association between the treatment differences on SVR and the clinical outcome.

An alternate approach to regression coefficients or correlation coefficient to decide whether an outcome is a good surrogate outcome (using the first approach) is using the models to predict the clinical outcome from the surrogate outcome (9). This approach uses a “take-one-out” approach to predict the treatment effect of the intervention on the clinical outcome in one study based on the observed treatment effect of the intervention on the surrogate outcome and meta-analytical estimate of the correlation between the surrogate outcome and clinical outcome in the remaining studies (9).

However, this approach is unlikely to be feasible in our study, since the long-term results of direct-acting antivirals are unlikely to be available from randomised clinical trials. In observational studies, the reason for receiving (or not receiving) treatment with direct-acting antivirals may be correlated with the clinical outcome. For example, in one study, the reasons for not receiving direct-acting antivirals included multiple comorbidities, low health literacy, restricted access to hospitals, nursing home residence, and old age (18), all of which are associated with increased risk of death. This means that confounding could be a major reason for an observed association between no treatment and mortality in the observational studies. Metaregression is a statistical method that uses study-level characteristics to provide information of whether the treatment effects are affected by the characteristic and is primarily used to explore statistical heterogeneity in meta-analyses (19). By using confounding factors as the characteristics, it is theoretically possible to calculate the meta-analytical treatment effects across studies adjusting for the differences in the confounders, for example, the differences in age or proportion of people with major co-morbidities between the studies. However, it is unlikely that the adjusted treatment effects of direct-acting antivirals on SVR and clinical outcome are calculated using the same confounding factors across studies; therefore, it is likely that information on some confounding factors are missing from some studies. This means it is unlikely that we will be able to use a metaregression approach to calculate the treatment effects of direct-acting antivirals on clinical outcome from the treatment effects on SVR adjusting for the values of the potential confounders. Furthermore, such a metaregression approach to adjust for study-level characteristics can lead to ecological bias (19,20). Therefore, we have used the approach of using the regression coefficients and correlation coefficient to achieve our research objective.

With our chosen second approach of causal mediation analysis, we will use the lower 95% CI of the proportion of not overlapping 0.5 as the criterion for indicating that SVR is a good surrogate outcome. This is an arbitrary decision but based on the rationale that SVR should at least explain the majority of the effect of direct-acting antivirals on the clinical outcome, if SVR is to be used as the primary objective of treatment or the primary outcome in a clinical trial. Since the 95% CI of the proportion is presented, it is possible for an individual or an organisation to use their own criterion for interpreting the information.

As indicated earlier in the discussion, there are possible confounding factors explaining the decision to treat (or not to treat a person with direct-acting antivirals). Therefore, we will analyse the data separately between randomised clinical trials and observational studies.

## Contributions

Kurinchi Gurusamy wrote the first draft. Christian Gluud commented and improved the draft.

---

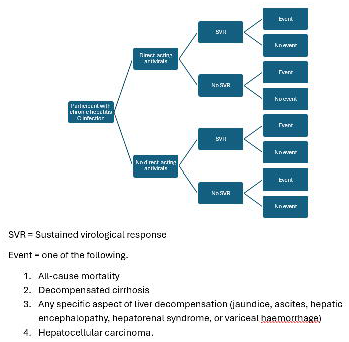

